# Dementia risk factors and cognitive decline: Results from the 2023 Behavioral Risk Factor Surveillance System (BRFSS)

**DOI:** 10.1101/2025.01.27.25321203

**Authors:** Mary L. Adams

## Abstract

**Background:** The potentially modifiable risk factors of obesity, diabetes, hypertension, smoking, physical inactivity, depression, hearing impairment, and excessive alcohol consumption, have been found to be associated with cognitive decline and dementia.

**Objective:** To study the association between subjective cognitive decline (SCD) among survey respondents and the 8 risk factors compared with other potential risks.

**Methods:** Using 2023 Behavioral Risk Factor Surveillance data from 155,112 respondents ages 45+ in 35 states with data on SCD the associations between the 8 dementia risk factors, separately and in a composite measure, plus other demographic and health measures were studied using Stata.

**Results:** SCD rates were 17.2% overall ranging from 11.0% to 23.9% across states while 81.9% of adults 45+ reported any of the 8 dementia risk factors. Adults with each of the risk factors had higher SCD rates compared with those without the risk factor, with SCD rates ranging from 8.4% for adults with 0 risks, 19.2% for any of the 8 risks, and 45.3% for those with 5 or more. Logistic regression confirmed most results with adjusted odds ratios (AORs) up to 6.0 for 5 or more risk factors, >3.1 for current long COVID and >1.2 for low education. Obesity was not confirmed as a separate risk and highest AOR for age was 1.2 for ages 75+ with 8 risk factors entered separately.

**Conclusions:** Results suggest that prevention efforts for SCD, which can be a step toward dementia, should focus on 8 common potentially modifiable risk factors plus long COVID.

## Introduction

Several potentially modifiable risk factors for dementia have been identified with more added as they are discovered. The current list of 12 includes obesity, diabetes, hypertension, smoking, physical inactivity, depression, low education, hearing impairment, excessive alcohol consumption, low social contact, traumatic brain injury, and air pollution which were estimated to account for up to 40% of all Alzheimer’s Disease (AD) cases worldwide [1]. That study updated an earlier one [2] that had estimated up to half of AD cases could be attributed to the first 7 of the 12 RFs. That earlier study [2] was among the evidence the Alzheimer’s Association considered in its 2014 report [3] that concluded that there was strong evidence for the first 5 risk factors and less support for cognitive inactivity/low education and depression as increasing the risk of cognitive decline. Cognitive impairment has also been studied as an early step in progression to AD which is the most common form of dementia [4,5]. More recently, similarities have been noted between cognitive issues and COVID-19; for example, post-COVID cognitive dysfunction (PCCD) has been described [6] as a condition in which patients who had long COVID exhibit subsequent cognitive impairment that cannot be explained by an alternate diagnosis. More evidence of the similarities between COVID and cognitive issues comes from studies showing COVID-19 is a risk factor for AD [7,8] and that patients with AD are at increased risk of severe complications and death from COVID [9].

The main objective of this current study was to study the association between the dementia risk factors of obesity, diabetes, hypertension, smoking, physical inactivity, depression, hearing impairment, and excessive alcohol consumption, plus low education and cognitive decline as measured on the same survey among adults ages 45 and older. Separate risk factors and a composite measure including all 8 potentially modifiable risks plus a demographic measure of not finishing high school will be used in the study. Because of evidence suggesting dementia and long COVID may be associated [7-9] current long COVID will be included among the measures studied. The 35 states which included the optional Cognitive Decline module on their 2023 BRFSS will be used for the study.

## Methods

The study used publicly available 2023 telephone survey data from 155,112 adults ages 45 and older in the 35 states that asked the cognitive decline module on the ongoing Behavioral Risk Factor Surveillance System (BRFSS). Data and questionnaires are available on the Centers for Disease Control and Prevention (CDC) website [10]. Files from different survey versions (6 from version 2 and 2 from version 3) were combined with the common version per CDC instructions [11] to create the final data set. Data were already weighted to adjust for the probability of selection and to reflect the adult population of each state by gender, age, race/ethnicity, education level, marital status, home ownership, regions within states, and telephone source. The median response rate for the 35 states for land line and cell phone surveys combined was 44.9% [10], ranging from 33.9% in NY to 58.4% in ND.

Measures: Subjective cognitive decline (SCD) was defined as a “yes” response to this question only asked of respondents ages 45 and older “During the past 12 months, have you experienced difficulties with thinking or memory that are happening more often or are getting worse?” A related disability question asked on all federal surveys [12, 13] was also asked: “Because of a physical, mental, or emotional condition, do you have serious difficulty concentrating, remembering, or making decisions?” The latter question appears to be an acceptable measure for cognitive impairment (CI) but not cognitive decline because it lacks a time frame. Other measures included age, race/ethnicity, gender, education, employment, census region (Northeast, Midwest, South and West), state of residence, and long COVID, which was defined as having a positive test for COVID and currently having symptoms lasting 3 months or longer that were not present prior to COVID-19 [10]. In addition, 8 of the 12 risk factors shown to be associated with dementia [1] (obesity, diabetes, depression, physical inactivity {no leisure time physical activity}, hypertension, current smoking, excessive drinking {binge or heavy drinking as defined separately for male and female respondents}, and hearing impairment were included as “dementia risks”. Other measures included general health status (fair or poor vs good or better), frequent (14 or more days in past 30) of poor mental or physical health or activity limitation.

Data on traumatic brain injury, low social contact, and air pollution [1] were not available. Low education was included as less than a high school education in the demographic measure of education. The 8 dementia risks (excluding low education) were included in a composite measure to check for dose response gradients with increasing numbers of risk factors.

Analysis: Stata version 18.0 (StataCorp LLC, College Station TX) was used to account for the complex sample design of the BRFSS in unadjusted analysis and controlled for the listed factors in logistic regression. The survey measures used to describe the survey design were _psu and _ststr, weight was _llcpwt or the survey version weight; linearized variance estimation was selected, with the option to center at the grand mean for strata with a single sampling unit. Missing values were excluded from analysis of that variable. Variables to be included in logistic regression models were selected from results of univariate analysis. Results for logistic regression are presented separately for models using the 8 separate dementia risk factors or the composite measure.

## Results

The prevalence of SCD for adults ages 45 and older (Table 1) was 17.2% overall (95% confidence interval 16.7-17.6) and only slightly higher among ages 75+ (20.4%) compared with those 45-75 years (16.1-16.8). Highest rates for SCD were found for those reporting CI (65.2%), 5 or more dementia risks (45.3%), frequent mental distress (FMD) (44.0%) or the single dementia risk factor of depression (38.9%) and the lowest rate of 8.4% was among respondents 45+ reporting none of the 8 dementia risk factors. Results for the 8 separate risk factors are listed in Table 1 from highest to lowest prevalence among those with SCD. Not shown are results indicating that 81.9% of study adults age 45+ reported any of the 8 dementia risk factors with results across states ranging from 82.2% in CO to 89.1% in MS and 29.9% reported ≥3 with a range from 20.1% to 42.8% across the same states. Current long COVID (not shown) was reported by 7.0% (6.7-7.3) of all adults ages 45+ with rates decreasing from 9.1% for adults 45-54 years to 4.1% for those age 65+ years. Among adults with long COVID 37.1% reported SCD. Cognitive decline rates were also higher among women, American Indian/AK natives and adults of multiple races, those who had not completed high school, or were unable to work or not working. Rates were lower among Asians and those in Northeast and Midwest regions compared with the West and South (Table 1). Measures of health status indicated that adults with SCD were more likely than those without SCD to report poor general health and 14+ days of both poor mental health and activity limitations in the past month.

**Table 1.**
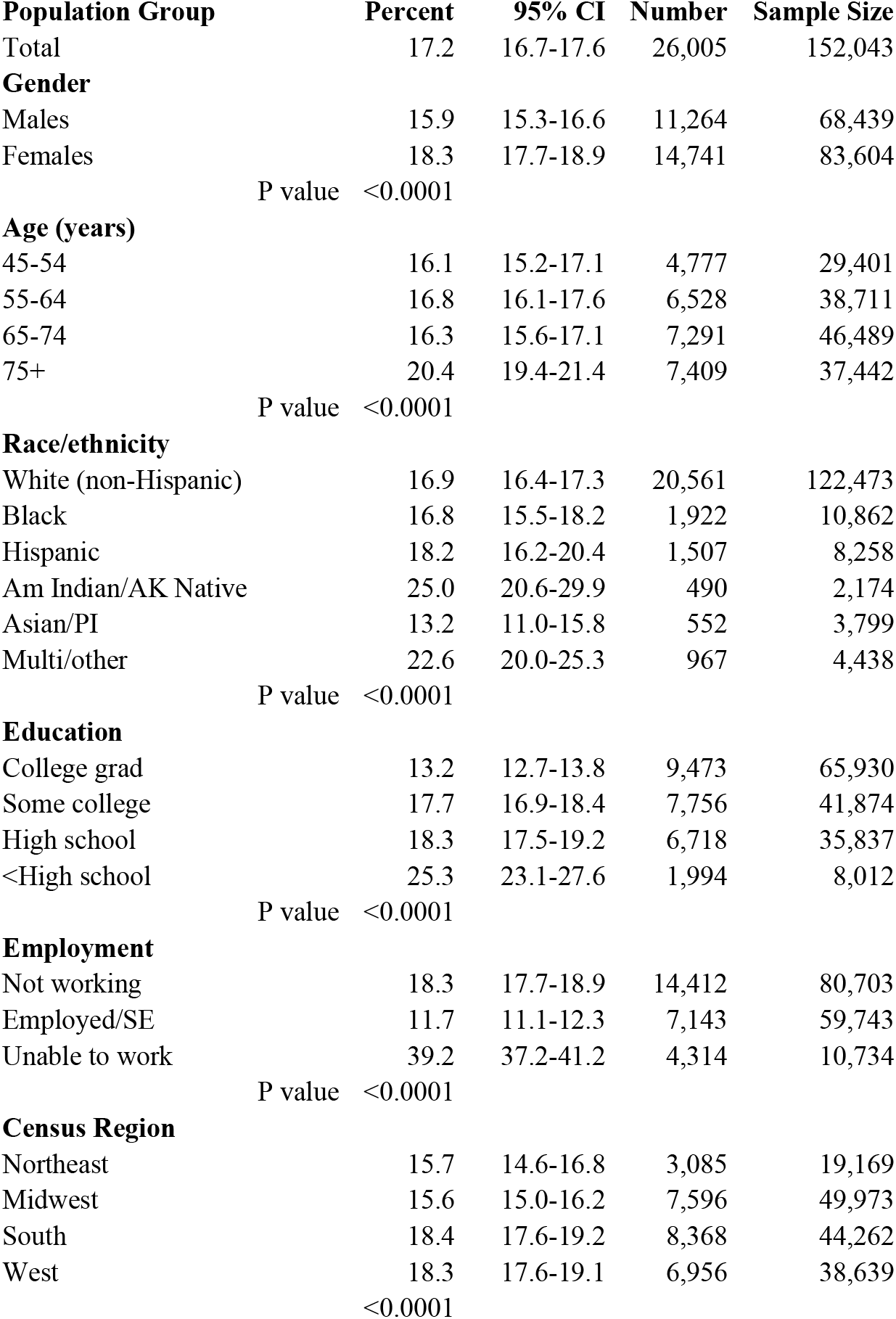
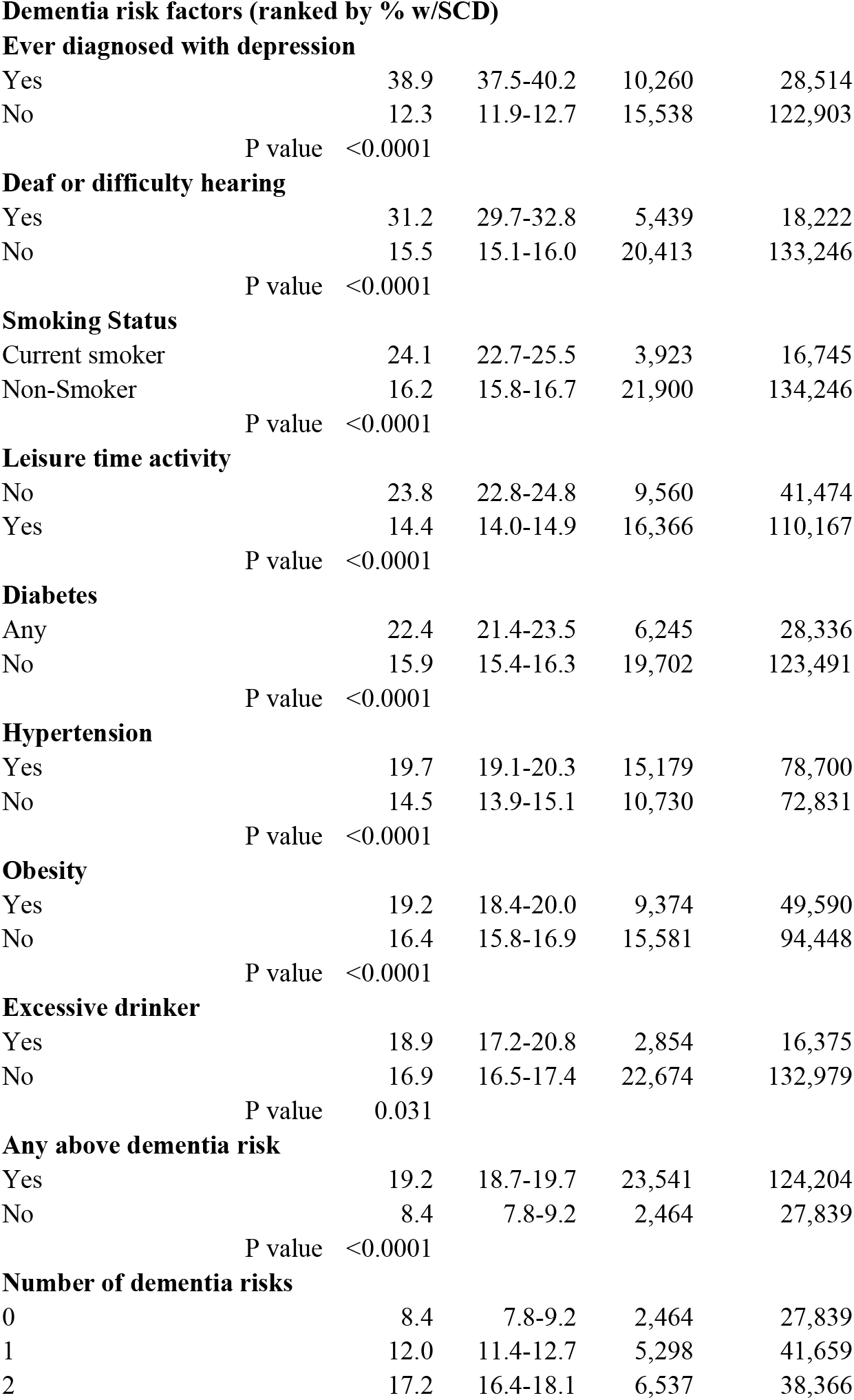
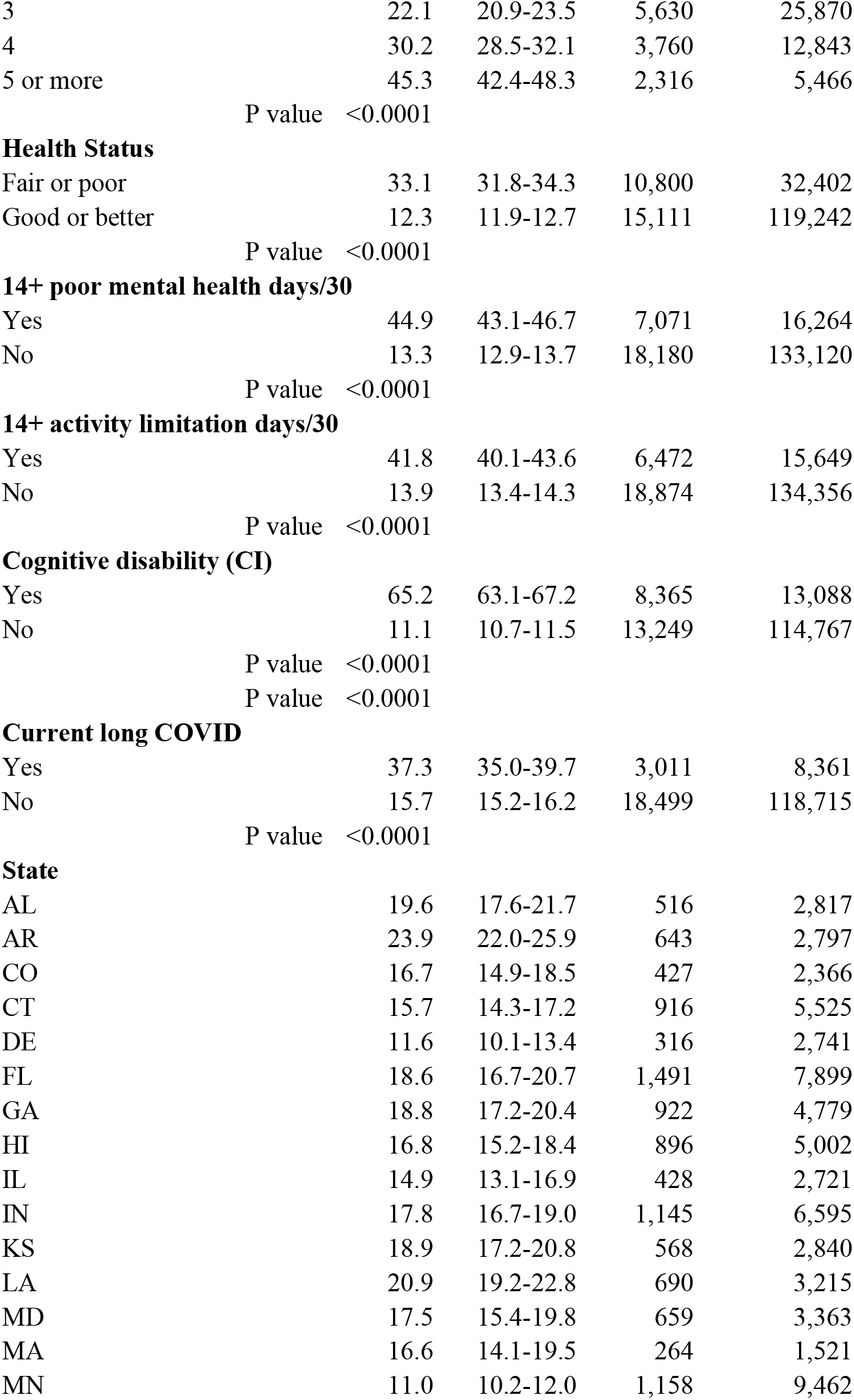
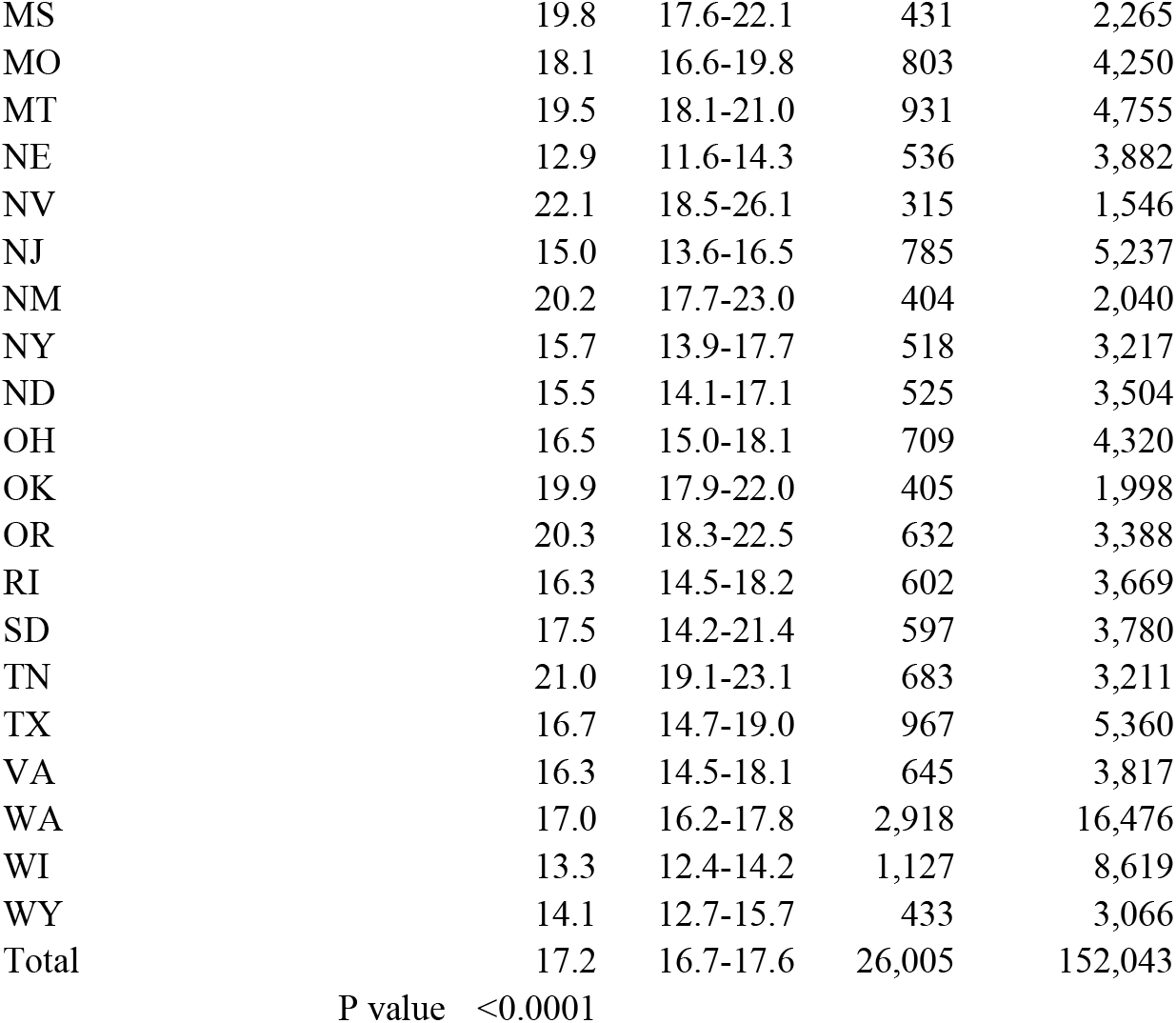
Subjective Cognitive Decline ages 45+, 35 states, 2023 Behavioral Risk Factor Surveillance System (BRFSS)

Results for logistic regression found that if the disability measure of CI was included in the models, 7 or 8 states were dropped from analysis due to collinearity, reducing the sample size considerably. Because this did not happen when the CI measure was excluded from the regression model, analysis was limited to models without CI. Other methods may be used in future to better measure association of CI and SCD. There was a clear indication that adults with SCD were much more likely to report CI (i.e. cognitive issues that were not worsening) compared with adults the same age without SCD.

Logistic regression results (Tables 2 & 3) show slightly different results for the model with the 8 separate dementia risk factors compared with the composite measure, in both cases including all listed measures in the model. When the composite measure was included in the model a distinct dose-response reaction was noted with increasing number of risks and the highest adjusted odds ratio (AOR) was 6.0 for ≥5 risk factors. For the model with the 8 separate risk factors the highest AOR was for the separate measure of a depression diagnosis with AOR=3.7. Current long COVID had an AOR > 3.1 in models with either the separate risk factors or the composite measure while the models had AORs of between 1.2 and 1.3 for not completing high school.

**Table 2.**
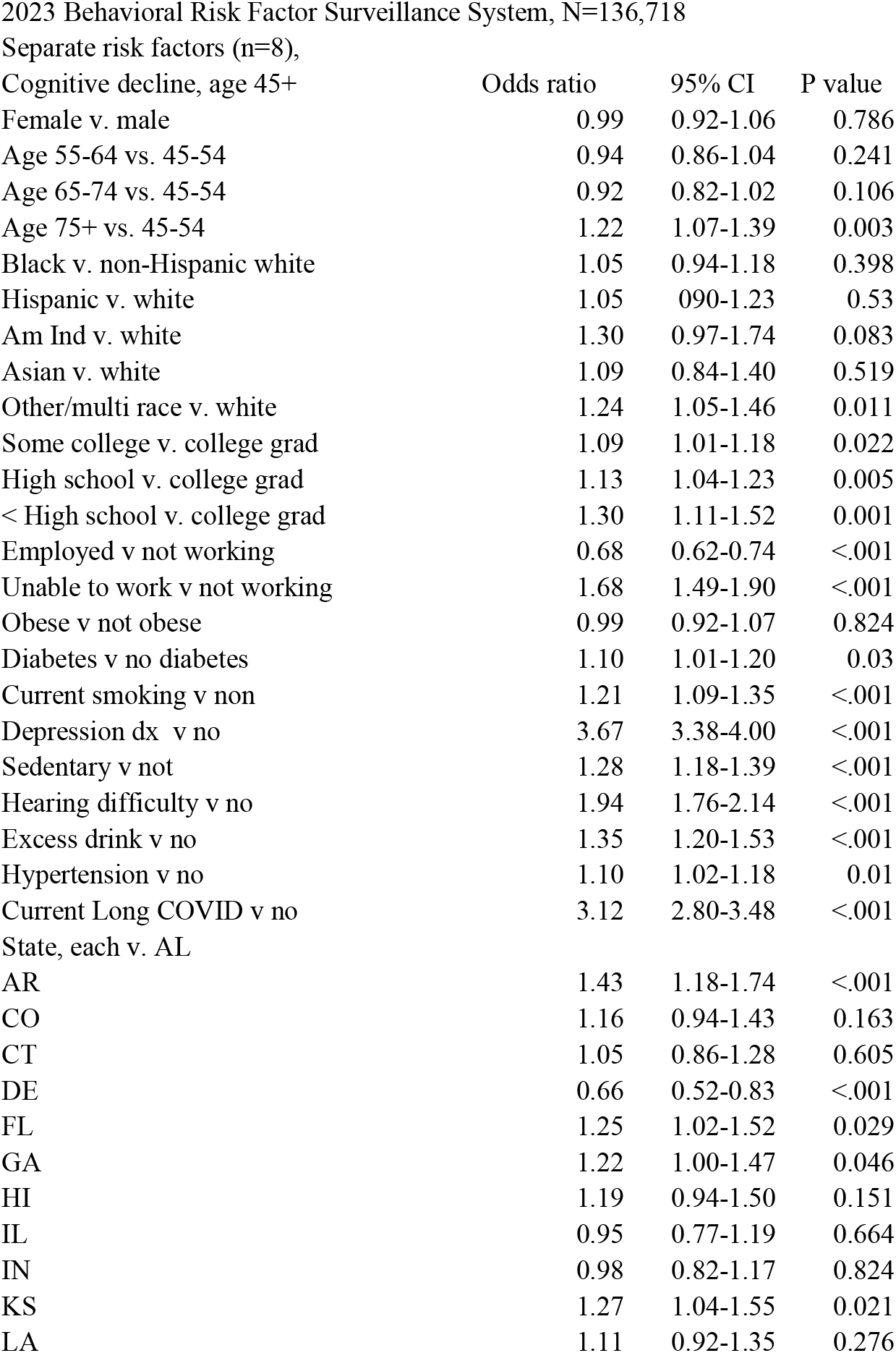
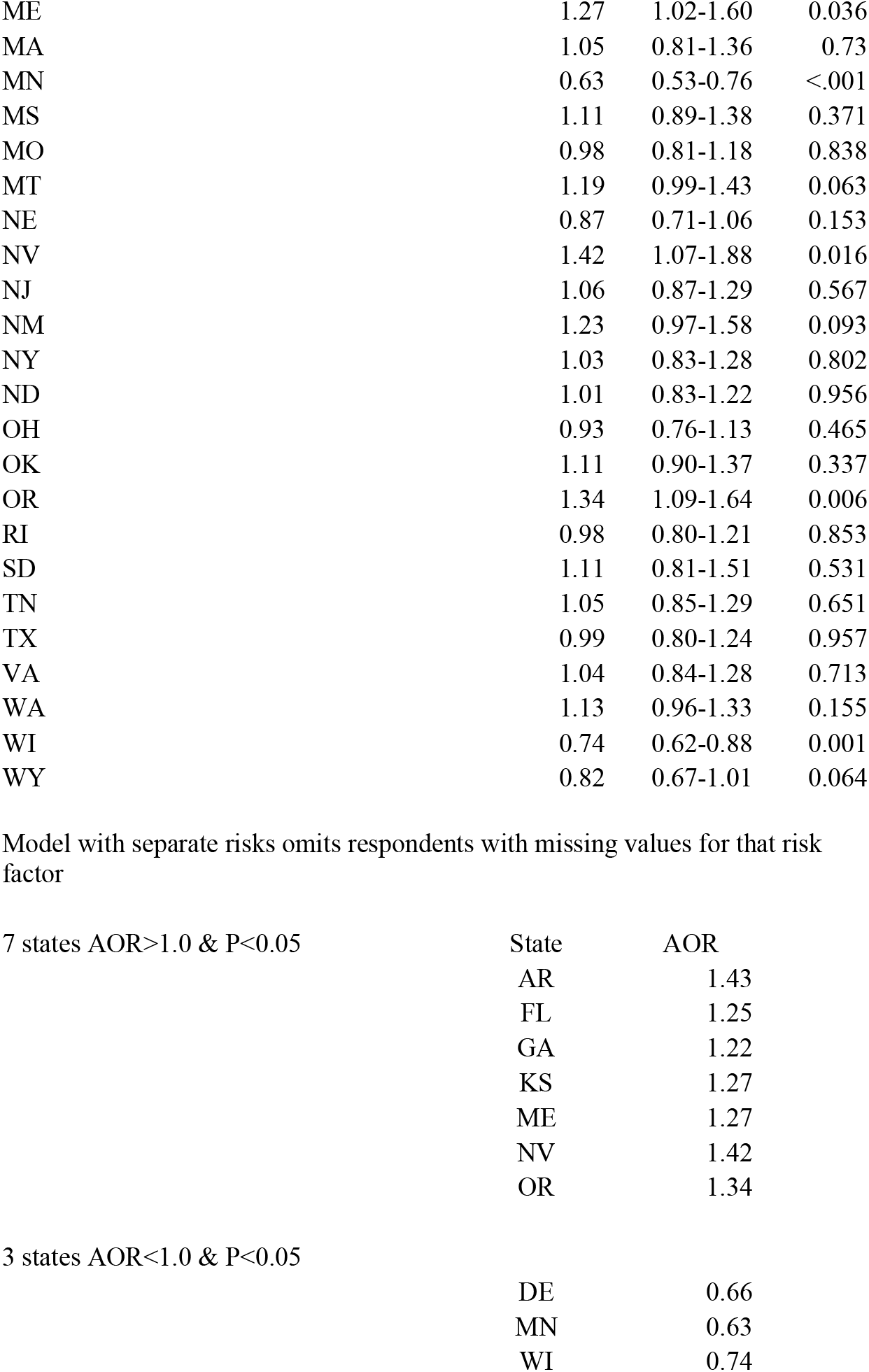
Logistic regression with outcome cognitive decline, adults age 45+

| Cognitive decline, age 45+ | Odds ratio | 95% CI | P value |
| --- | --- | --- | --- |
| Female v. male | 0.99 | 0.92-1.06 | 0.786 |
| Age 55-64 vs. 45-54 | 0.94 | 0.86-1.04 | 0.241 |
| Age 65-74 vs. 45-54 | 0.92 | 0.82-1.02 | 0.106 |
| Age 75+ vs. 45-54 | 1.22 | 1.07-1.39 | 0.003 |
| Black v. non-Hispanic white | 1.05 | 0.94-1.18 | 0.398 |
| Hispanic v. white | 1.05 | 0.90-1.23 | 0.53 |
| Am Ind v. white | 1.30 | 0.97-1.74 | 0.083 |
| Asian v. white | 1.09 | 0.84-1.40 | 0.519 |
| Other/multi race v. white | 1.24 | 1.05-1.46 | 0.011 |
| Some college v. college grad | 1.09 | 1.01-1.18 | 0.022 |
| High school v. college grad | 1.13 | 1.04-1.23 | 0.005 |
| < High school v. college grad | 1.30 | 1.11-1.52 | 0.001 |
| Employed v not working | 0.68 | 0.62-0.74 | <.001 |
| Unable to work v not working | 1.68 | 1.49-1.90 | <.001 |
| Obese v not obese | 0.99 | 0.92-1.07 | 0.824 |
| Diabetes v no diabetes | 1.10 | 1.01-1.20 | 0.03 |
| Current smoking v non | 1.21 | 1.09-1.35 | <.001 |
| Depression dx v no | 3.67 | 3.38-4.00 | <.001 |
| Sedentary v not | 1.28 | 1.18-1.39 | <.001 |
| Hearing difficulty v no | 1.94 | 1.76-2.14 | <.001 |
| Excess drink v no | 1.35 | 1.20-1.53 | <.001 |
| Hypertension v no | 1.10 | 1.02-1.18 | 0.01 |
| Current Long COVID v no | 3.12 | 2.80-3.48 | <.001 |
| State, each v. AL |  |  |  |
| AR | 1.43 | 1.18-1.74 | <.001 |
| CO | 1.16 | 0.94-1.43 | 0.163 |
| CT | 1.05 | 0.86-1.28 | 0.605 |
| DE | 0.66 | 0.52-0.83 | <.001 |
| FL | 1.25 | 1.02-1.52 | 0.029 |
| GA | 1.22 | 1.00-1.47 | 0.046 |
| HI | 1.19 | 0.94-1.50 | 0.151 |
| IL | 0.95 | 0.77-1.19 | 0.664 |
| IN | 0.98 | 0.82-1.17 | 0.824 |
| KS | 1.27 | 1.04-1.55 | 0.021 |
| LA | 1.11 | 0.92-1.35 | 0.276 |
| ME | 1.27 | 1.02-1.60 | 0.036 |
| MA | 1.05 | 0.81-1.36 | 0.73 |
| MN | 0.63 | 0.53-0.76 | <.001 |
| MS | 1.11 | 0.89-1.38 | 0.371 |
| MO | 0.98 | 0.81-1.18 | 0.838 |
| MT | 1.19 | 0.99-1.43 | 0.063 |
| NE | 0.87 | 0.71-1.06 | 0.153 |
| NV | 1.42 | 1.07-1.88 | 0.016 |
| NJ | 1.06 | 0.87-1.29 | 0.567 |
| NM | 1.23 | 0.97-1.58 | 0.093 |
| NY | 1.03 | 0.83-1.28 | 0.802 |
| ND | 1.01 | 0.83-1.22 | 0.956 |
| OH | 0.93 | 0.76-1.13 | 0.465 |
| OK | 1.11 | 0.90-1.37 | 0.337 |
| OR | 1.34 | 1.09-1.64 | 0.006 |
| RI | 0.98 | 0.80-1.21 | 0.853 |
| SD | 1.11 | 0.81-1.51 | 0.531 |
| TN | 1.05 | 0.85-1.29 | 0.651 |
| TX | 0.99 | 0.80-1.24 | 0.957 |
| VA | 1.04 | 0.84-1.28 | 0.713 |
| WA | 1.13 | 0.96-1.33 | 0.155 |
| WI | 0.74 | 0.62-0.88 | 0.001 |
| WY | 0.82 | 0.67-1.01 | 0.064 |

Model with separate risks omits respondents with missing values for that risk factor 7 states AOR>1.0 & P<0.05
| State | AOR |
| --- | --- |
| AR | 1.43 |
| FL | 1.25 |
| GA | 1.22 |
| KS | 1.27 |
| ME | 1.27 |
| NV | 1.42 |
| OR | 1.34 |

3 states AOR<1.0 & P<0.05
|  |  |
| --- | --- |
| DE | 0.66 |
| MN | 0.63 |
| WI | 0.74 |

**Table 3.**
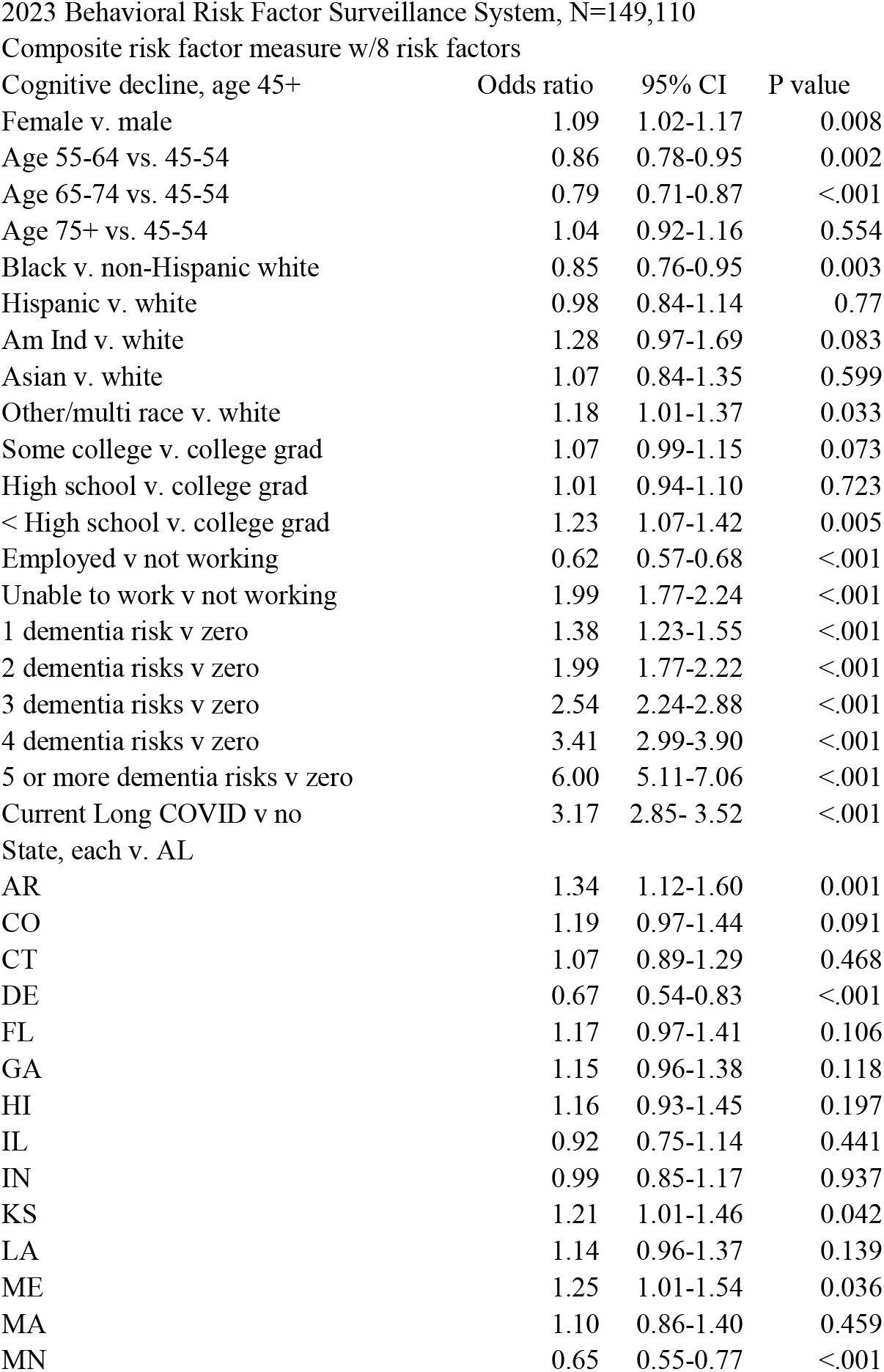
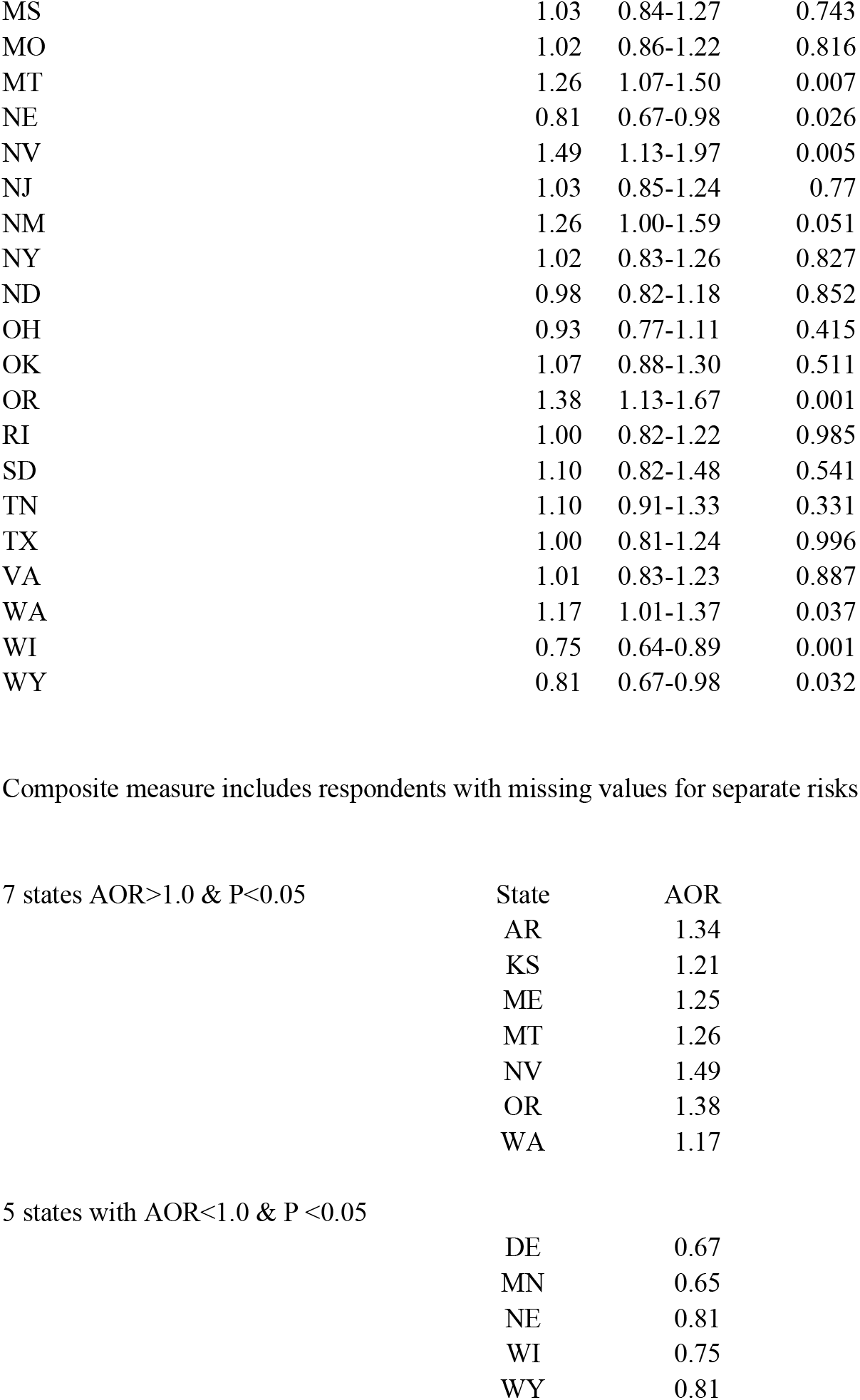
Logistic regression with outcome=cognitive decline, adults age 45+

| Cognitive decline, age 45+ | Odds ratio | 95% CI | P value |
| --- | --- | --- | --- |
| Female v. male | 1.09 | 1.02-1.17 | 0.008 |
| Age 55-64 vs. 45-54 | 0.86 | 0.78-0.95 | 0.002 |
| Age 65-74 vs. 45-54 | 0.79 | 0.71-0.87 | <.001 |
| Age 75+ vs. 45-54 | 1.04 | 0.92-1.16 | 0.554 |
| Black v. non-Hispanic white | 0.85 | 0.76-0.95 | 0.003 |
| Hispanic v. white | 0.98 | 0.84-1.14 | 0.77 |
| Am Ind v. white | 1.28 | 0.97-1.69 | 0.083 |
| Asian v. white | 1.07 | 0.84-1.35 | 0.599 |
| Other/multi race v. white | 1.18 | 1.01-1.37 | 0.033 |
| Some college v. college grad | 1.07 | 0.99-1.15 | 0.073 |
| High school v. college grad | 1.01 | 0.94-1.10 | 0.723 |
| < High school v. college grad | 1.23 | 1.07-1.42 | 0.005 |
| Employed v not working | 0.62 | 0.57-0.68 | <.001 |
| Unable to work v not working | 1.99 | 1.77-2.24 | <.001 |
| 1 dementia risk v zero | 1.38 | 1.23-1.55 | <.001 |
| 2 dementia risks v zero | 1.99 | 1.77-2.22 | <.001 |
| 3 dementia risks v zero | 2.54 | 2.24-2.88 | <.001 |
| 4 dementia risks v zero | 3.41 | 2.99-3.90 | <.001 |
| 5 or more dementia risks v zero | 6.00 | 5.11-7.06 | <.001 |
| Current Long COVID v no | 3.17 | 2.85- 3.52 | <.001 |
| State, each v. AL |  |  |  |
| AR | 1.34 | 1.12-1.60 | 0.001 |
| CO | 1.19 | 0.97-1.44 | 0.091 |
| CT | 1.07 | 0.89-1.29 | 0.468 |
| DE | 0.67 | 0.54-0.83 | <.001 |
| FL | 1.17 | 0.97-1.41 | 0.106 |
| GA | 1.15 | 0.96-1.38 | 0.118 |
| HI | 1.16 | 0.93-1.45 | 0.197 |
| IL | 0.92 | 0.75-1.14 | 0.441 |
| IN | 0.99 | 0.85-1.17 | 0.937 |
| KS | 1.21 | 1.01-1.46 | 0.042 |
| LA | 1.14 | 0.96-1.37 | 0.139 |
| ME | 1.25 | 1.01-1.54 | 0.036 |
| MA | 1.10 | 0.86-1.40 | 0.459 |
| MN | 0.65 | 0.55-0.77 | <.001 |
| MS | 1.03 | 0.84-1.27 | 0.743 |
| MO | 1.02 | 0.86-1.22 | 0.816 |
| MT | 1.26 | 1.07-1.50 | 0.007 |
| NE | 0.81 | 0.67-0.98 | 0.026 |
| NV | 1.49 | 1.13-1.97 | 0.005 |
| NJ | 1.03 | 0.85-1.24 | 0.77 |
| NM | 1.26 | 1.00-1.59 | 0.051 |
| NY | 1.02 | 0.83-1.26 | 0.827 |
| ND | 0.98 | 0.82-1.18 | 0.852 |
| OH | 0.93 | 0.77-1.11 | 0.415 |
| OK | 1.07 | 0.88-1.30 | 0.511 |
| OR | 1.38 | 1.13-1.67 | 0.001 |
| RI | 1.00 | 0.82-1.22 | 0.985 |
| SD | 1.10 | 0.82-1.48 | 0.541 |
| TN | 1.10 | 0.91-1.33 | 0.331 |
| TX | 1.00 | 0.81-1.24 | 0.996 |
| VA | 1.01 | 0.83-1.23 | 0.887 |
| WA | 1.17 | 1.01-1.37 | 0.037 |
| WI | 0.75 | 0.64-0.89 | 0.001 |
| WY | 0.81 | 0.67-0.98 | 0.032 |

Composite measure includes respondents with missing values for separate risks 7 states AOR>1.0 & P<0.05
| State | AOR |
| --- | --- |
| AR | 1.34 |
| KS | 1.21 |
| ME | 1.25 |
| MT | 1.26 |
| NV | 1.49 |
| OR | 1.38 |
| WA | 1.17 |

5 states with AOR<1.0 & P <0.05
|  |  |
| --- | --- |
| DE | 0.67 |
| MN | 0.65 |
| NE | 0.81 |
| WI | 0.75 |
| WY | 0.81 |

Controlled for all factors in the model, each model found 7 states with AORs >1.0 although they were not the same states, and either 3 or 5 states with AORs <1.0.

## Discussion

This study shows that 8 of the 12 dementia risk factors shown to account for 40% of dementia and AD cases worldwide [1] also increase rates of cognitive decline for adults 45+ in survey data. Cognitive decline among adults able to respond to a survey suggests these adults may be at an early stage in possible progression to dementia. This study confirms that all 8 dementia risk factors increased unadjusted rates of SCD in adults ages 45+ and all except obesity increased odds ratios in logistic regression models. In the model with the composite risk factor measure that included all 8 risks, the AOR for adults with ≥5 risk factors = 6.0. While not included as one of the dementia risk factors, not finishing high school also increased SCD rates in unadjusted and adjusted results. These results are all consistent with earlier studies [1-3]. What appears new are the findings that 1) over 80% of surveyed adults aged 45+ reported at least one of the 8 dementia risk factors available for the study (obesity, diabetes, depression, physical inactivity, hypertension, excessive drinking, current smoking, and hearing impairment, and 2) current long COVID appeared to be a significant risk factor for SCD with an AOR >3.1. An earlier study in 21 states [14] that defined 6 dementia risk factors as obesity, diabetes, depression, inactivity, current smoking and hypertension found 77.3% of adults ages 45 and older reported any of the 6 factors. That study included 232 adults with dementia and 9,769 with cognitive decline and found dose response gradients for more risk factors for both outcomes in unadjusted and adjusted results. For those few with all 6 risk factors the highest AOR was 11.2 for cognitive decline and over 100 for adults with dementia (with a wide confidence interval). Thus, results from this current study are consistent with earlier results which included dementia along with cognitive decline and included 6 of the 8 risk factors used here.

In this current study, where 81% of all adults reported any of the 8 dementia factors, 29.3% reported 3 or more. In the older study [14] also limited to adults ≥45 years and using 6 of these 8 risk factors, about 25% of all adults reported 3 or more dementia risk factors. These results add key information because the study which estimated 40% of AD cases could be attributed to 12 dementia risk factors [10] had very limited data that included all or most of the risk factors together. These new results suggest that addressing multiple risk factors (i.e. 3 or more together) could be a key issue in planning prevention programs. Results also illustrate the challenge of decreasing future AD cases if the magnitude of an early step in the progression to AD can be increased based on the presence of these common risk factors. While this study notes which of the risk factors individually are more common, further study is needed to determine which of the risk factors are most likely to be grouped together. The differences in logistic regression results between states when controlled for so many demographic factors suggest further study to determine what other factors might be contributing and how they might be mitigated.

These results for long COVID appear to support findings that COVID-19 can be a risk factor for cognitive decline or AD [7,8] or that survivors of COVID are at increased risk of developing AD [9]. The rate of long COVID in 2023 of 7.0% among study adults ages 45+ is only slightly lower than the 7.4% rate found for all age adults in 2022 [15]. While that study used a slightly different definition of long COVID and age group, in both studies adults with long COVID represented one in five adults with a positive COVID test suggesting little or no decline in long COVID rates between 2022 and 2023. Along with determining which of the dementia risk factors are most likely to be grouped together, it will be important to see how results might change if long COVID is considered a dementia risk factor and included in the composite measure. These results, taken together with results from other studies, suggest the possibility that COVID-19 may remain a threat to our healthcare systems for the foreseeable future due to the potential for increased AD cases.

A very recent paper estimates the lifetime risk of dementia after age 55 at 42% with higher rates in women, Blacks and APOE ε4 carriers [16]. The number of US adults estimated to develop dementia each year was projected to increase from about 500,000 in 2020 to approximately 1 million in 2060. Those estimates are unlikely to have considered any possible contributions due to COVID or long COVID as found in this study.

A meta-analysis of US data that included all 12 of the dementia risk factors estimated population attributable fractions (PAF) overall and for different race and ethnicities [17]. That study estimated that 41.0% (95% CI, 22.7%-55.9%) of dementia cases were associated with these 12 risk factors, very similar to the 40% noted earlier [1] with higher estimates for Blacks and Hispanics than for Whites or Asians. That study estimated that a 15% decrease in each risk factor would reduce dementia prevalence by an estimated 7.3% (95% CI, 3.7%-10.9%). The greatest attributable fractions of dementia cases were contributed by hypertension (PAF, 20.2%; 95% CI, 6.3%-34.4%), obesity (PAF, 20.9%; 95% CI, 13.0%-28.8%), and physical inactivity (PAF, 20.1%; 95% CI, 9.1%-29.6%). A more recent study [18] included more countries and addressed later-life dementia (adults ages 60 years or older). They found PAFs for individual risk factors were generally higher in low- and middle-income countries compared with higher income ones like the US.

What results of these studies have in common is the demonstrated need to reduce the substantial and growing burden of dementia by targeting modifiable risk factors. Delaying dementia onset in the population seems to be the best strategy to curb the projected increase in dementia cases. Top priority should be to reduce the prevalence of obesity, hypertension, and physical inactivity, which are currently associated with the largest fraction of dementia cases in the US. Efforts to reduce racial and ethnic disparities should continue. In all cases, emphasis should be placed on interventions shown to be successful.

## Limitations

A major limitation results from the requirement that survey respondents need to be able to respond to a telephone survey and as SCD progresses, this ability is likely affected. Once a randomly selected respondent is deemed unable to answer the survey, they are no longer included in the BRFSS and the entire household is dropped from the sample. Another BRFSS study that included non-respondents in households with respondents found that some measures of cognitive decline were under-reported by as much as 70% when only respondent data was included [19]. A related limitation is that only households are included so nursing homes and assisted living facilities are excluded and of course, anyone under age 45 was also excluded.

Some study limitations are common to any studies using survey data: 1) results are self-reported and not based on an actual test or diagnosis; 2) survey results can’t distinguish cause and effect; and 3) only 35 states had survey data on cognitive decline. The study’s strengths are that the data that are available for the 35 states are population based and demonstrate the variation in results among the states included. As noted above, having multiple risk factors available from the same data set allows study of associations between risk factors that might be important when planning interventions.

## Conclusion

Despite the limitations, these results indicate these 8 common potentially modifiable risk factors should be addressed in efforts to prevent SCD and/or slow development of dementia. Results also suggest that attention should be paid to the possible role of long COVID in increasing risk of SCD and dementia. In this study those factors were shown to have a much greater association with SCD than education or age.

## Data availability

All data used in the study are available on the CDC website at: https://www.cdc.gov/brfss/annual_data/annual_2023.html

